# IL-15 and sMAdCAM: Novel roles in COVID-19 pathogenesis

**DOI:** 10.1101/2021.03.25.21254215

**Authors:** Amit Kumar Singh, Nandini Kasarpalkar, Shilpa Bhowmick, Gaurav Paradkar, Mayur Talreja, Karan Shah, Abhishek Tiwari, Harsha Palav, Snehal Kaginkar, Rajiv Kulkarni, Ashwini Patil, Varsha Kalsurkar, Sachee Agrawal, Jayanthi Shastri, Rajesh Dere, Ramesh Bharmal, Smita D. Mahale, Vikrant M. Bhor, Vainav Patel

## Abstract

Immune cell dysregulation and lymphopenia characterize COVID-19 pathology in moderate to severe disease. While underlying inflammatory factors have been extensively studied, homeostatic and mucosal migratory signatures remain largely unexplored as causative factors. In this study we evaluated the association of circulating IL-6, soluble mucosal addressin cell adhesion molecule (sMAdCAM) and IL-15 with cellular dysfunction characterizing mild and hypoxemic stages of COVID-19. A cohort of SARS-CoV-2 infected individuals (n=125) at various stages of disease progression together with healthy controls (n=16) were recruited from COVID Care Centres (CCCs) across Mumbai, India. Multiparametric flow cytometry was used to perform in-depth immune subset characterization and to measure plasma IL-6 levels. sMAdCAM, IL-15 levels were quantified using ELISA. Distinct depletion profiles, with relative sparing of CD8 effector memory and CD4+ regulatory T cells was observed in hypoxemic disease within the lymphocyte compartment. An apparent increase in the frequency of intermediate monocytes characterized both mild as well as hypoxemic disease. IL-6 levels inversely correlated with those of sMAdCAM and both markers showed converse associations with observed lympho-depletion suggesting opposing roles in pathogenesis. Interestingly, IL-15, a key cytokine involved in lymphocyte activation and homeostasis, was detected in symptomatic individuals but not in healthy controls or asymptomatic cases. Further, negative association of plasma IL-15 with depleted T, B and NK subsets suggested a compensatory production of this cytokine in response to the profound lymphopenia. Finally, higher levels of plasma IL-15 and IL-6, but not sMAdCAM, were associated with longer duration of hospitalization.

## Introduction

The Severe acute respiratory syndrome coronavirus 2 (SARS-CoV-2) pandemic continues to pose a global health crisis in spite of ongoing interventions such as vaccination (1, 2). Pathology of COVID-19 displays varied clinical manifestations ranging from no symptoms to critical systemic disease (3–5). Hypoxemia is a key signature that discriminates between mild and moderate to severe disease(6, 7). The role of lymphopenia as a defining cellular immune correlate of moderate to severe disease has been well established (8). However, underlying immune homeostatic mechanisms that might contribute to this phenotype remain largely unexplored (9, 10). Understanding and identifying such relationships would help to guide therapeutic efforts and vaccine strategies to ensure optimal disease management of COVID-19. In this study we evaluated the contribution of key inflammatory, cellular homeostatic and mucosal migratory markers in distinct stages of COVID-19 pathogenesis. Our results highlight associations of IL-6, IL-15 and sMAdCAM with lymphopenia together with a heretofore undescribed role for detectable plasma IL-15 as marker associated with symptomatic progression.

## Results

### Demographic and Clinical characteristics of study participants

A total of 125 admitted hospital patients, laboratory-confirmed positive for SARS-CoV-2 infection by quantitative RT-PCR of throat and nasal swab samples were recruited for the study. According to the clinical management protocol for COVID-19 (Version 3) issued by Ministry of Health and Family Welfare, Government of India, patients were recruited into different groups based on clinical severity(7). The patients without evidence of breathlessness or hypoxia were classified into Mild group. Mild patients were further segregated into Asymptomatic Mild (AS; n=47) and Symptomatic Mild (SM; n=57) where SMs presented mild symptoms like fever, cough, sore throat, headache etc. Patients with oxygen saturation (SpO_2_) of < 93% on room air who required oxygen support to correct hypoxemia, were classified as having Moderate (MD; n=10) COVID-19, whereas patients with oxygen saturation (SpO_2_) of < 90% on room air were considered as having Severe (SV; n=11) COVID-19. In addition, a total of 16 healthy participants with no apparent history of COVID-19 and seronegative (SN) by Rapid antibody test, were also recruited as controls. The demographic and clinical characteristics of all the study participants is summarised in Table 1.

**Table 1.** Abridged clinical data of study participants

|  | Asymptomatic<br>mild | Symptomatic<br>mild | Moderate | Severe |
| --- | --- | --- | --- | --- |
| Number of<br>Participants | 47 | 57 | 10 | 11 |
| Age<sup>a</sup> (years) | 39 (18-75) | 37 (18-71) | 64 (47-75) | 66 (31-81) |
| Gender (M/F) | 33/14 | 41/16 | 6/4 | 9/2 |
| SpO<sub>2</sub> levels | - | - | <93% | <90% |
| Duration of<br>Hospitalisation<sup>a</sup><br>(days) | 8 (1-41) | 7 (1-41) | 12 (2-27) | 18 (8-38) |

**Footnote:** <sup>a</sup> Data are expressed as the median (range). SpO<sub>2</sub> levels at the time of recruitment are reflected in the table

### Lymphopenia with relative sparing of Treg and CD8+ effector memory subsets define hypoxemic COVID-19 progression

The absolute counts of total lymphocytes and T cells (both CD4+ and CD8+) in MD and SV groups were significantly decreased compared to AM, SM and SN (Supplementary figures S1 and S2A). Additionally, SV group also showed significant depletion of total lymphocytes compared to MDs. Similar depletion profiles (in MD and SV groups) were observed for both T (CD4+ and CD8+) and non T (B and NK) cell compartments. However, no significant difference was observed among mild patients (AM & SM) and compared to SN (Supplementary figure S2B-G). No apparent difference in the frequency of these lymphocyte subsets was observed except for a significant decrease for total T lymphocytes and a significant increase for total non-T lymphocytes in both MD and SV compared to other groups (Supplementary figure S2H-M). This suggested a pan lymphopenic phenotype that was observed in hypoxemic (MD and SV) individuals. Additionally, to evaluate the impact of lymphopenia on T cell compartment, distribution of CD4+ and CD8+ T cell subsets was also evaluated(11). Based on the surface expression of CD45RA and CCR7, CD4+ and CD8+ T cells were differentiated into Naïve (T_N;_ CD45RA+CCR7+), Central memory (T_CM;_ CD45RA-CCR7+), Effector memory (T_EM;_ CD45RA-CCR7-) and Terminally differentiated (T_TD;_ CD45RA+CCR7-) T cells (Supplementary figure S3). While both MD and SV groups had significant depletion of absolute count for all CD4+ T cell subsets (Figure 1A), our findings do not support preferential depletion of any particular subsets in the CD4+ T cell compartment (Figure 1B). In CD8+ T cell compartment, comparison of absolute counts revealed profound depletion of all subsets in hypoxemic (MD and SV) individuals (Figure 1C). In contrast to the CD4+ T cell compartment however, we observed relatively lower frequency of naïve (CD8+ T_N_) and concomitant elevation of effector memory (CD8+ T_EM_) frequency (Figure 1D). This reflected a distinct depletion profile for CD8+ T cells with relative sparing of effector memory cells.

**Figure 1.**
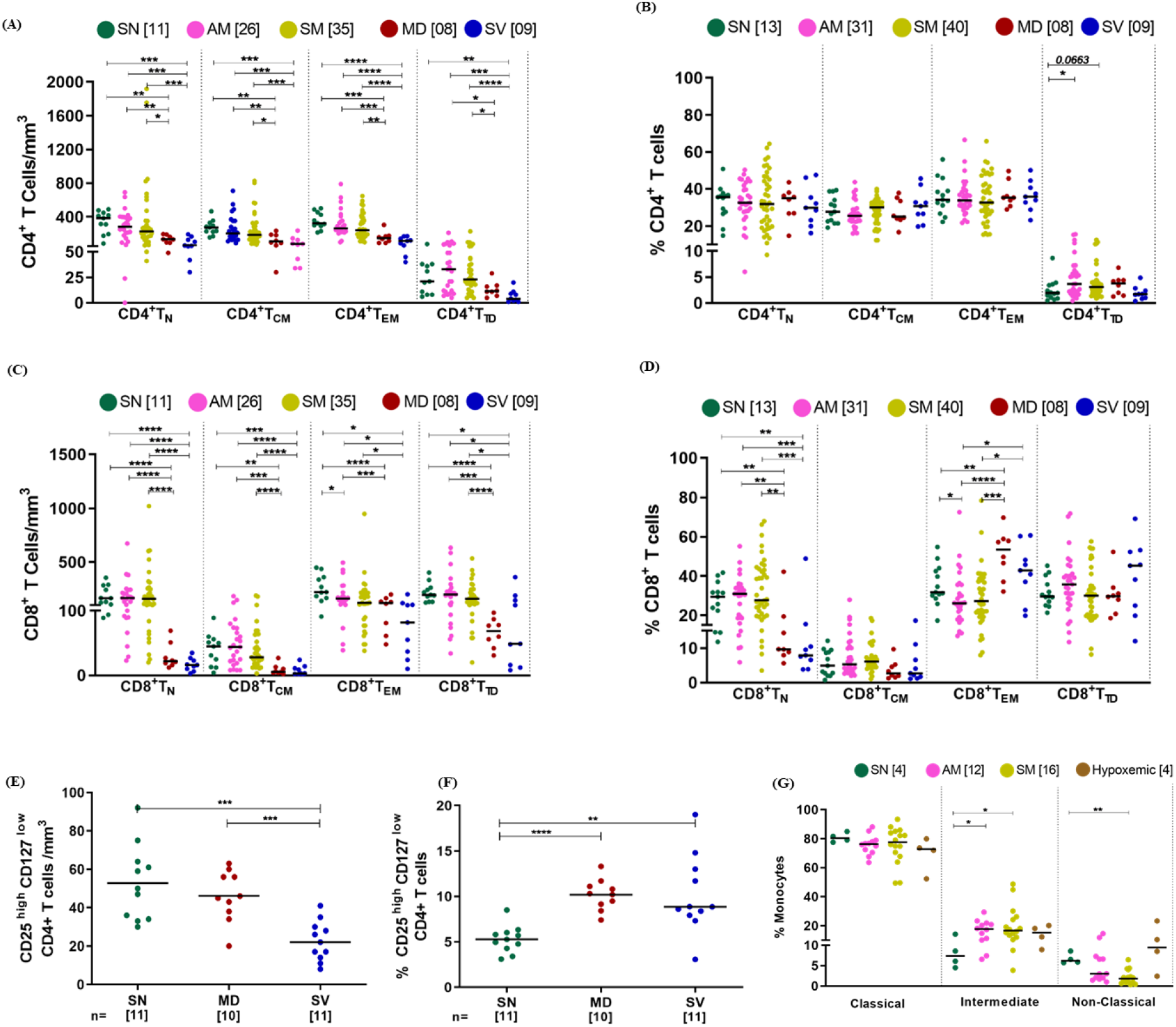
Distribution of different immune subsets among study participants. **(A-B)** Variation across seronegative (SN), asymptomatic mild (AM), symptomatic mild (SM), moderate (MD) and severe (SV) groups in absolute count and frequency of CD4+ T cells subsets (N= Naïve, CM= central memory, EM= Effector Memory and TD= Terminally differentiated), respectively. (**C-D)** Variation across different study groups in absolute count and frequency of CD8+ T cells subsets, respectively. (**E-F)** Variation across different groups in Tregs count (E) and frequency(F). (**G)** Variation in frequency of monocyte subsets across different study groups. Statistical significance was calculated by Mann-Whitney U-test; *, p < 0.05; **, p < 0.01, ***, p<0.001 ; and ****, p<0.001.

Interestingly, while memory subsets of CD4+ T cell compartment seemed to be equally depleted in hypoxemic individuals, we report here depletion in Treg count that was exacerbated in the SV group (Figure 1E). Also, elevated frequency of CD4+ T regulatory cells in hypoxemic individuals indicated selective preservation of this subset (Figure 1F). When the non-lymphocyte monocyte compartment was evaluated we observed an elevated frequency of the intermediate (CD14+CD16++) subset that defined both mild (AM, SM) and hypoxemic patients (Figure 1G).

### Contribution of sMAdCAM and IL-6 to COVID-19 associated lymphopenia

To assess the contribution of known soluble inflammatory cytokines and mucosal migration markers to the observed cellular immune signatures we undertook the evaluation of circulating IL-6 and sMAdCAM levels in our cohort. As expected, we observed elevated IL-6 levels, with the highest being observed in hypoxemic individuals, in plasma of all groups compared to seronegative controls (Figure 2A). Additionally, extending our previously reported results(12) obtained with sMAdCAM in mild infection, we observed a progressive decline in these levels across mild as well as hypoxemic patients that seemed converse to the pattern observed for IL-6 (Figure 2B). Indeed, the levels of these 2 markers were significantly negatively correlated (Figure 2C). Further, it was interesting to note that neither in the case of IL-6 or sMAdCAM was it possible to discriminate between AM and SM individuals. Next, correlation analysis was undertaken to delineate putative relationships between the aforementioned soluble markers and cellular subsets described above (Supplementary Figure S4). Intriguingly and reflective of their apparently divergent relationship, both IL-6 and sMAdCAM showed significant opposing correlations with absolute counts of lymphocytes, T cells (CD4+ and CD8+), B cells and NK cells supporting their clear, albeit, opposing roles in COVID-19 associated lymphopenia (Figure 2 D-I). We also noted a unique negative correlation of IL-6 with Treg counts together with a heretofore unreported positive correlation of sMAdCAM levels with CD8+ effector memory T cell counts (Supplementary Figure S5A-B). Elevated LPS levels, associated with microbial translocation and severe COVID-19 disease were observed in hypoxemic individuals (Supplementary Figure S5C). Furthermore, and possibly related to altered monocyte frequencies observed ex vivo (Figure 1G), a negative correlation of sMAdCAM levels occurred with frequencies of intermediate monocytes (Supplementary Figure S5D). Gut pathology associated markers LPS and sMAdCAM were poorly correlated with only the latter exhibiting major correlations to lymphopenia.

**Figure 2.**
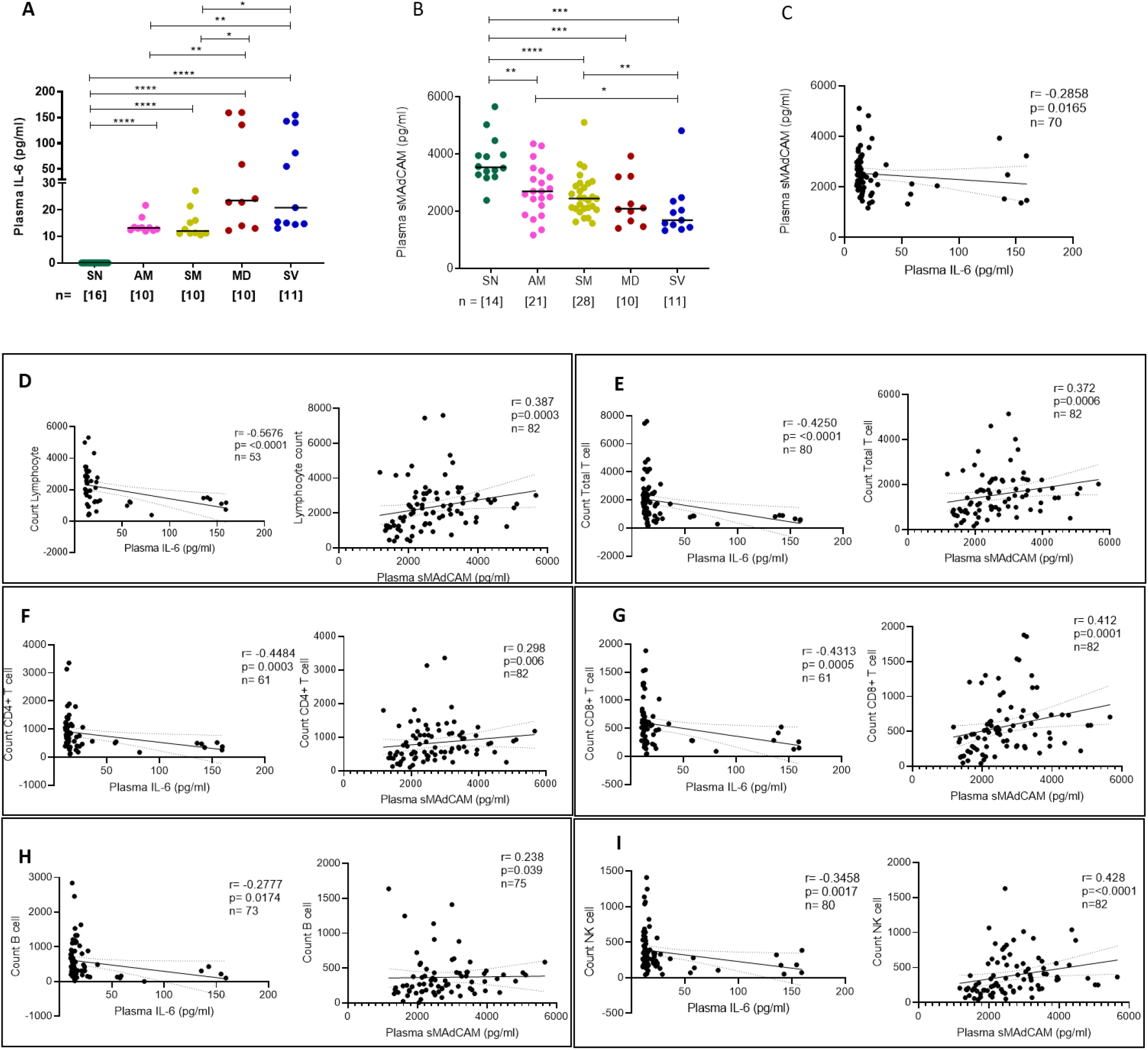
Evaluation of IL-6 and sMAdCAM along with correlation of cellular immune subsets in study participants. Variation in levels of (A) IL-6 and (B) sMAdCAM in seronegative(SN), asymptomatic mild (AM), symptomatic mild (SM), moderate (MD) and severe (SV) groups. Relationship between (C) IL-6 and sMAdCAM and their association with different cellular subsets (D) Lymphocyte count (E) Total T cell count (F) CD4+ T cell count (G) CD8+ T cell count (H) B cell count (I) NK cell count. Statistical significance was calculated by Mann-Whitney U-test; *, p < 0.05; **, p < 0.01, ***, p<0.001 ; and ****, p<0.001. Correlation analysis was performed using non parametric Spearman Rank Correlation test.

### Plasma IL-15 levels discriminate symptomatic and asymptomatic mild COVID-19

Having observed distinct depletion profiles of CD4+ and CD8+ T cells, the relative sparing of CD8+ effector memory T cells and non T cell lymphopenia (B and NK cells) we surmised that IL-15, a major homeostatic and activation cytokine for these subsets needed to be evaluated in COVID-19 pathogenesis. We observed elevated levels of circulating IL-15 in lymphopenic, hypoxemic individuals and remarkably, as shown in Figure 3A, in symptomatic mild but not asymptomatic patients. All hypoxemic individuals had detectable levels of IL-15 followed by 70% of individuals with symptomatic infection and only 18% (2 out of 11) with asymptomatic infection (Figure 3B). Indeed, of the 2 asymptomatic individuals that had detectable levels of IL-15 at sampling, one later progressed to hypoxemia. No individuals within the SN control group had detectable levels of plasma IL-15. Underlying a strong role for this cytokine, possibly related to homeostatic repopulation in lymphopenia we observed significant negative correlations between depleted subsets and circulating IL-15 levels across all groups of COVID-19 affected individuals (Figure 3C-H and Supplementary Figure S4). Also, as observed for IL-6, sMAdCAM levels were negatively correlated with those of IL-15 (Figure 3I).

**Figure 3.**
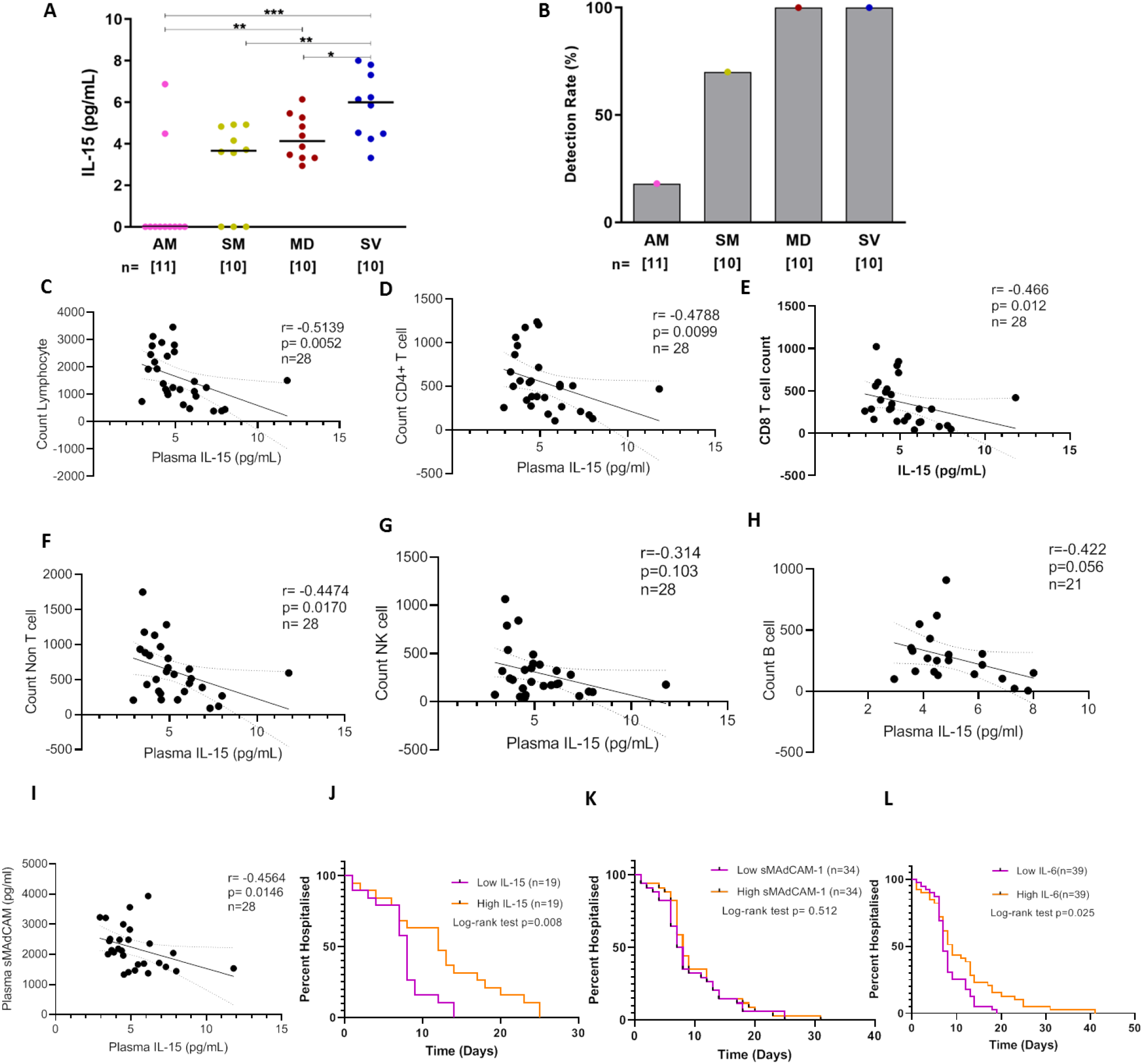
Distribution of IL-15 and its correlation with immune cells among study participants. (A) Variation in levels of IL-15 in asymptomatic mild (AM), symptomatic mild (SM), moderate (MD) and severe (SV) groups. (B) Responder rate. Association of IL-15 and with different cellular subsets (C) Lymphocyte count (D) Total T cell count (E) CD4+ T cell count (F) CD8+ T cell count (G) B cell count (H) NK cell count (I) Soluble MAdCAM. (J-L) Kaplan Meier estimates of hospital discharge following estimation of plasma (J) IL-15 (K) sMAdCAM and (L) IL-6. Individuals were categorized into a low and a high group on the basis of median estimated levels of either IL-15, sMAdCAM or IL-6. Statistical significance was calculated by Mann-Whitney U-test; *, p < 0.05; **, p < 0.01, ***, p<0.001; and ****, p<0.001. Correlation analysis was performed using non parametric Spearman Rank Correlation test.

Finally, in a rudimentary analysis, we evaluated the possible prognostic value of our single determinations of IL-15 together with IL-6 and sMAdCAM in predicting duration of hospitalization independently. For this analysis, days of hospitalization post sampling, including those from individuals with undetectable values for these markers (considered as 0) were plotted as shown in Figure 3J-L. Proportional hazards analysis revealed significantly longer hospitalization for individuals having IL-15 and IL-6 values, but not sMAdCAM values, in the upper 50 percent of our data sets. This value was above >4.15 pg/ml of IL-15 and >14.29 pg/ml for IL-6.

## Discussion

In this study we describe cellular immune signatures characterizing mild (asymptomatic as well as symptomatic) and hypoxemic (moderate and severe) COVID-19 disease. We further elucidate the role of sMAdCAM, an important mucosal and lymphocyte migration marker and IL-15, a cytokine regulating leukocyte activation and homeostasis, in the observed cellular immune signatures. Our results clearly illustrate, as has been well described (13–16), leukopenic profiles only in hypoxemic COVID-19 affected individuals. Further, in-depth analysis of T cell subsets revealed distinct depletion profiles in operation within CD4+ and CD8+ T cell compartments. While the CD4+ T cell compartment did not show any preferential depletion across naïve and memory subsets, we report here an asymmetric depletion profile in hypoxemic COVID-19 infection characterized by drastic depletion of naïve CD8+ T cells with relative sparing of the CD8+ effector memory T cell subset. These results extend the limited data available for T cell subsets in a stratified manner to include distinct stages of COVID-19 pathogenesis (10, 17). The CD4+ T regulatory cell subset represents a critical part of the response to viral infections (18, 19) that may play both protective as well as exacerbatory roles in COVID-19 pathogenesis. Our analysis revealed an incremental decrease in absolute count of this subset from moderate to severe pathogenesis compared to healthy controls. However, both stages of hypoxemic infection showed a relative preservation of this subset evidenced through elevated frequencies compared to healthy controls. As has been reported very recently this elevated frequency may represent suppressive (protective) as well as inflammatory roles played by these cells, markedly higher, in severe disease (20). Our study also evaluated the non-lymphocyte compartment in terms of monocytes and their profiles in mild and hypoxemic COVID-19 disease. This compartment has been known to modulate inflammatory responses to a variety of viral infections (21, 22). Interestingly, as opposed to the T cell compartment and when compared with healthy seronegative individuals, a pan-disease signature was delineated within monocytes comprising of elevated frequencies of intermediate monocytes. Also, concomitant decrease in frequency of non-classical monocytes was observed in mild infection irrespective of symptoms.

In an effort to understand factors influencing the observed cellular depletion and dysfunction in our cohort we evaluated plasma markers that could influence inflammation, mucosal leukocyte migration and homeostasis. Extending and strengthening our previous results on the converse relationship between sMAdCAM and IL-6 observed in non-hypoxemic COVID-19 progression (12), we report a similar and inversely correlated relationship between sMAdCAM and IL-6 levels across mild and hypoxemic stages of COVID-19 disease. Furthermore, for the first time, we show that this converse relationship extends to opposing correlations with observed cellular immune signatures defining COVID-19 pathogenesis. Notably, elevated circulating LPS (a marker for gut inflammation), in the context of SARS-CoV-2 infection, has only been reported for severe and ICU patients (9). In our study we also observed increased plasma LPS levels in hypoxemic individuals compared to seronegative controls as opposed to our previous work with a cohort of mild COVID-19 (12). Importantly, there was no correlation between circulating LPS levels and either sMAdCAM or IL-6 levels. These results suggest that the altered trajectory of sMAdCAM levels is less related to gut persistence mediated inflammation and more to altered mucosal leukocyte migration. Taken together, our results show that IL-6 and sMAdCAM may contribute towards disparate aspects of disease progression where circulating levels of the former are more closely related to inflammatory sequalae and development of anti-viral B cell responses (class switching IL-6 reference) and those of the latter with restoration of mucosal homeostasis of leukocytes. Indeed, we have recently highlighted a role for the modulation of sMAdCAM and expression of its receptor integrin alpha-4 beta-7 present on monocytes, lymphocytes in the context of HIV pathogenesis (23).

While most immune markers of pathogenesis in COVID-19 have been reported to be associated with mild, moderate and severe disease, a marker that clearly discriminates between symptomatic and asymptomatic pathogenesis remains elusive. Our study is the first to demonstrate the presence of plasma IL-15 as such a marker where detection of this cytokine is clearly segregated based on the symptomatic status of patients with incrementally higher levels detected in mild symptomatic to hypoxemic individuals. In fact, of the 2 (out of 11) mild asymptomatic individuals that had detectable levels of this cytokine, one was found retrospectively to have progressed to a hypoxemic stage. Also, negative correlation of plasma IL-15 levels with lymphopenic profiles (T, B, NK) are indicative of its homeostatic restorative production (24–26) in the face of systemic depletion of these subsets. Circulating IL-15 levels have been implicated as a contributory factor to hospitalization time, disease severity and mortality in some settings (27, 28). Indeed, a preliminary analysis in our cohort, with respect to days of hospitalization supported this finding. Our study further demonstrates the utility of using it as an early biomarker predicting progression to symptomatic and severe disease.

## Conclusion

In summation, our results delineate a role for inflammatory, homeostatic and migratory immune mediators in COVID-19 associated lymphopenia, of which, plasma IL-15 detection may be an early prognostic marker.

## Methods

### Study population, setting, and data collection

Sixteen seronegative controls and a total of 125 in-patients individuals were recruited, following informed consent, for the study from the COVID Care Centres associated with BYL Nair hospital and T N medical college, Municipal Corporation of Greater Mumbai (MCGM), Mumbai following approval of institutional ethics committees. We obtained demographic data, clinical history at presentation, and laboratory results during admission.

Blood samples for the study were handled in accordance with ICMR guidelines for biosafety. 1-3ml of whole blood was collected in EDTA vacutainers. Aliquots of whole blood were processed for absolute cell count and immunophenotyping as described below. Plasma was separated by centrifugation at 400 g for 10 minutes. IgG and IgM antibodies against SARS-CoV-2 were detected in fresh plasma samples using Rapid test from Voxpress (Voxtur Bio LTD, India) and Chemiluminescence immunoassay (CLIA) directed against SARS-CoV-2 anti-NC IgG. Remaining plasma samples were aliquoted and stored at −80^0^ C until batch analysis of cytokines, LPS (Lipopolysaccharide) and Soluble MAdCAM (sMAdCAM).

### Assay for absolute count of T cells, B cells and NK cells

BD multitest 6 colour TBNK reagent and BD Trucount™ tubes were used to enumerate absolute count of T cells, CD4 T cells, CD8 T cells, B cells, and Natural killer (NK) cells using 50 μl of fresh EDTA stabilised blood following stain/lyse/no-wash protocol. Data acquisition was performed on BD FACS Aria Fusion (SORP) Flow cytometer (BD Biosciences) where 5000 events gated on beads count were acquired. Data analysis was carried out using FlowJo software (BD biosciences)

### Immunophenotyping

For phenotypic characterization, immunostaining of 200 μl of fresh peripheral whole blood with following fluorescently labelled monoclonal antibodies, anti-CD3 (Clone:SK7), anti-CD4 (Clone: RPA-T4), anti-CD8 (Clone: SK1), anti-CD25 (Clone: M-A251), anti-CD127 (Clone: HIL-7R-M21), anti-CCR7 (Clone: 150503), anti-CD45RA (Clone: HI100), anti-CD14 (Clone: M5E2) and anti-16 (Clone: 3G8) was performed using stain/lyse/wash protocol as described earlier (11). Data acquisition was performed on BD FACS Aria Fusion (SORP) flow cytometer (BD Biosciences) where at least 100,000 events in lymphocyte scatter gate were acquired. Data analysis was carried out using FlowJo software.

### Cytometric Bead Array

Flow based Cytometric bead array (Human Th1/Th2/Th17 cytokine kit, BD Biosciences) was used to quantify cytokine levels in plasma of study participants as per manufacturers’ protocol.

### ELISA

Plasma IL-15, sMAdCAM-1 and LPS levels were quantified using Human IL-15 DuoSet ELISA kit (R&D Systems), Human MAdCAM-1 DuoSet ELISA kit (R&D Systems) and Human Lipopolysaccharides ELISA kit (MyBioSource) respectively, following manufacturers’ recommendations. The values below the level of detection were assigned as zero.

### Statistics

Statistical analysis was performed in GraphPad Prism 8 using non parametric tests. Statistical significance of differences between groups were assessed using Mann-Whitney U-test. Spearman’s rank-order correlation was used to analyse the association between participant attributes that had detectable values in our assays. Kaplan–Meier curves were compared using Log-rank (Mantel-Cox) test. Statistical significance was accepted at p<0.05.

## Data Availability

All data available for review upon request to corresponding authors.

## Author Contribution

Sample processing, assay running, data analysis and figure generation (Amit Kumar Singh, Nandini Kasarpalkar, Shilpa Bhowmick). Participant recruitment, sample processing and data analysis (Gaurav Paradkar, Mayur Talreja). Sample processing (Abhishek Tiwari, Harsha Palav, Snehal Kaginkar). Participant recruitment (Rajiv Kulkarni, Karan Shah). Sample collection (Ashwini Patil, Varsha Kalsurkar). Performed experiment and Data analysis (Sachee Agrawal, Jayanthi Shastri). Scholarly advice (Rajesh Dere, Ramesh Bharmal, Smita Mahale). Conceived study, analysed data, compiled results and wrote manuscript (Vikrant Bhor and Vainav Patel). All authors revised the manuscript and gave final approval for publication.

## Acknowledgement

We are extremely grateful to Director General, Indian Council of Medical Research (ICMR), Ministry for Health & Family Welfare, Government of India for encouraging and supporting with resources, the pursuit of important research questions relevant to COVID-19 pathogenesis. We would also like to thank Dr. Noora Pathan, Dr. Mahesh Chaurasia, Dr. Rasika Satpute and Dr. Dharmesh Balsarkar for facilitating recruitment of study participants and Mr. Omkar Arekar for the assistance provided in data entry. Also, none of this work would have been possible without the active participation and support of participants to whom we are grateful.

