## Supplementary data for "IL-15 and sMAdCAM: Novel roles in COVID-19 pathogenesis"

***Supplementary Material***

**Table S1. Study cohort characteristics**

| **Patient Id** | **Age** | **Sex** | **Comorbidities** | **Duration of Hospitalisation (days)** | **Ct values** | | **Antibody** | | **C.L.I.A** |
| --- | --- | --- | --- | --- | --- | --- | --- | --- | --- |
|  |  |  |  |  | ***ORF*** | ***E*** | **IgM** | **IgG** |  |
| X0209 | 60s | F | Diabetes mellitus, Hypertension | 12 | - | - | - | - | - |
| X0212 | 70s | M | Diabetes mellitus, Hypertension | 8 | 26.35 | 26.38 | + | + | 4.18 |
| X0214 | 60s | F | Hypertension | 5 | - | - | - | + | 1.56 |
| X0215 | 50s | M | No | 12 | 31.74 | 32.27 | - | + | - |
| X0217 | 70s | F | Hypertension | 15 | - | - | - | + | 4.35 |
| X0218 | 50s | M | No |  | - | - | + | + | 6.9 |
| X0221 | 60s | F | Hypertension | 2 | 26.88 | 26.7 | - | - | - |
| X0210 | 30s | F | Hypothroidism | 14 | 28.08 | 28.23 | - | - | - |
| X0213 | 60s | M | Hypertension | 34 | - | - | + | + | 6.55 |
| X0223 | 60s | M | No | 38 | 33 | 33 | - | + | 5.31 |
| X0225 | 50s | M | No | 27 | 35 | 34 | - | + | 6.97 |
| X0226 | 60s | M | No | 14 | - | - | + | + | 7.59 |
| X0233 | 60s | M | No | 13 | 24 | 24 | - | - | 2.3 |
| X0292 | 60s | F | No | 18 | - | - | - | + | 4.76 |
| X0293 | 40s | M | Diabetes mellitus | 10 | 35.88 | 37.09 | - | + | 3.46 |
| X0297 | 80s | M | No | 22 | 28.1 | 29.42 | - | + | 1.84 |
| X0299 | 60s | M | No | 28 | 25.56 | 26.05 | - | - | 1.29 |
| X0300 | 50s | M | No | 14 | 32.32 | 37.87 | - | + | 2.51 |
| X0362 | 60s | M | Diabetes mellitus, Hypertension | 11 | 33.93 | 37.25 | - | + | 5.52 |
| X0376 | 40s | M | Hypertension | 12 | 35.07 | 35.82 | - | + | 4.82 |
| X0379 | 70s | M | Diabetes mellitus | 26 | 33.36 | 36.1 | - | + | 5.24 |
| X0089 | 50s | M | Hypertension | 7 | 26.85 | 27.04 | - | - | 7 |
| X0090 | 20s | F | No | 6 | - | - | - | - | 1.51 |
| X0105 | 10s | M | No | 8 | 29.49 | 29.26 | - | - | - |
| X0106 | 20s | F | No | 7 | 23.02 | 22.97 | - | - | - |
| X0109 | 40s | F | Hypothroidism, Hypertension | 9 | 18.31 | 17.95 | - | - | N/A |
| X0110 | 40s | M | Diabetes mellitus | 7 | 24.89 | 24.8 | - | - | N/A |
| X0126 | 50s | M | No | 8 | - | - | - | - | - |
| X0127 | 20s | F | No | 8 | - | - | - | - | - |
| X0128 | 40s | M | No | 8 | 31.18 | 31.71 | - | - | - |
| X0130 | 30s | M | No | 12 | 24.01 | 23.98 | - | - | - |
| X0131 | 70s | M | No | 20 | 31.54 | 31.71 | - | - | N/A |
| X0132 | 20s | M | No | 14 | 17.71 | 17.65 | - | - | - |
| X0133 | 30s | F | No | 1 | - | - | - | + | N/A |
| X0136 | 30s | M | No | 12 | 19.86 | 19.33 | - | - | N/A |
| X0138 | 30s | M | No | 4 | - | - | - | - | - |
| X0142 | 40s | M | No | 8 | 33.56 | 35.6 | - | - | 7.43 |
| X0143 | 40s | M | No | 7 | 17.82 | 17.66 | - | - | - |
| X0145 | 20s | M | No | 14 | 23.66 | 23.36 | - | - | - |
| X0146 | 50s | M | No | 8 | 24.96 | 24.87 | - | - | - |
| X0150 | 20s | M | No | 7 | 29.61 | 29.75 | - | - | - |
| X0025 | 10s | M | No | 12 | 25.68 | 25.48 | N/A | N/A | N/A |
| X0026 | 40s | M | No | 9 | 25.77 | 25.62 | N/A | N/A | N/A |
| X0027 | 20s | M | No | 9 | 31.99 | 33.17 | N/A | N/A | N/A |
| X0028 | 50s | M | No | 10 | 25.51 | 25.26 | N/A | N/A | N/A |
| X0029 | 30s | F | No | 5 | 24.4 | 25.34 | N/A | N/A | N/A |
| X0030 | 30s | F | No | 5 | 29.85 | 30.19 | N/A | N/A | N/A |
| X0031 | 40s | F | No | 12 | 25.81 | 25.95 | N/A | N/A | N/A |
| X0032 | 20s | M | No | 7 | 25.23 | 25.2 | N/A | N/A | N/A |
| X0033 | 30s | F | No | 7 | 19.89 | 20.09 | N/A | N/A | N/A |
| X0045 | 40s | M | No | 14 | 25.52 | 25.12 | N/A | N/A | N/A |
| X0046 | 30s | M | No | 8 | 24.11 | 23.52 | N/A | N/A | N/A |
| X0048 | 40s | M | No | 5 | 20.51 | 20.11 | N/A | N/A | N/A |
| X0049 | 40s | F | Low BP | 7 | 29.25 | 29.57 | N/A | N/A | N/A |
| X0050 | 40s | M | Hypertension, Diabetes mellitus | 6 | 36.19 | 36.83 | N/A | N/A | N/A |
| X0054 | 20s | M | No | 7 | 18.22 | 18.06 | N/A | N/A | N/A |
| X0057 | 50s | M | No | 7 | 29.12 | 29.2 | N/A | N/A | N/A |
| X0058 | 30s | M | Hypertension | 7 | 24.02 | 23.68 | N/A | N/A | N/A |
| X0059 | 30s | M | No | 6 | 35.02 | 3716 | N/A | N/A | N/A |
| X0060 | 20s | M | No | 12 | 27.83 | 27.94 | N/A | N/A | N/A |
| X0062 | 40s | M | No | 6 | 22.95 | 22.36 | N/A | N/A | N/A |
| X0063 | 20s | M | No | 6 | 20.75 | 20.31 | N/A | N/A | N/A |
| X0065 | 30s | M | No | 7 | 25.48 | 25.2 | N/A | N/A | N/A |
| X0067 | 40s | M | No | 10 | 20.6 | 20.72 | N/A | N/A | N/A |
| X0068 | 30s | M | Diabetes mellitus | 6 | 26.1 | 26.36 | N/A | N/A | N/A |
| X0069 | 30s | F | No | 6 | 30.58 | 31.72 | N/A | N/A | N/A |
| X0070 | 30s | M | Hypertension | 6 | 20.04 | 2029 | N/A | N/A | N/A |
| X0071 | 40s | M | Hypertension | 6 | 23.38 | 23.51 | N/A | N/A | N/A |
| X0072 | 50s | M | Diabetes mellitus | 8 | - | - | N/A | N/A | N/A |
| X0073 | 40s | M | No |  | 32.37 | 34.5 | N/A | N/A | N/A |
| X0074 | 50s | M | No | 14 | 28.54 | 29.06 | N/A | N/A | N/A |
| X0075 | 40s | M | No | 5 | 21.14 | 21.41 | N/A | N/A | N/A |
| X0076 | 40s | M | No | 9 | 26.1 | 26.36 | N/A | N/A | N/A |
| X0077 | 30s | M | No | 6 | 18.6 | 18.91 | N/A | N/A | N/A |
| X0078 | 20s | M | No | 14 | 22.96 | 23 | N/A | N/A | N/A |
| X0079 | 30s | F | No | 7 | 32.57 | 34.93 | N/A | N/A | N/A |
| X0080 | 20s | M | No | 5 | 26.8 | 27.12 | N/A | N/A | N/A |
| X0081 | 40s | F | No | 7 | 25.19 | 25.5 | N/A | N/A | N/A |
| X0082 | 30s | M | No | 13 | 16.46 | 16.61 | - | - | - |
| X0083 | 20s | M | No | 2 | 28 | 28.31 | - | - | - |
| X0084 | 30s | M | No | 5 | 31.68 | 32.79 | - | + | 1.77 |
| X0085 | 30s | M | No | 13 | 19.67 | 1936 | - | - | - |
| X0086 | 30s | F | Hypertension | 6 | 27.72 | 28.07 | - | - | - |
| X0087 | 20s | M | No | 13 | 29.45 | 30.2 | - | - | - |
| X0088 | 30s | M | No | 41 | 29.86 | 30.68 | - | - | - |
| X0091 | 50s | F | Diabetes, Hypertension | 41 | 19.5 | 19.37 | - | - | - |
| X0093 | 50s | M | No | 9 | 19.58 | 19.71 | N/A | N/A | N/A |
| X0094 | 20s | M | No | 9 | 32.33 | 33.35 | N/A | N/A | N/A |
| X0095 | 50s | M | No | 9 | 22.84 | 23.04 | N/A | N/A | N/A |
| X0096 | 30s | M | No | 9 | 28.55 | 29.11 | N/A | N/A | N/A |
| X0098 | 20s | M | No | 1 | - | - | N/A | N/A | N/A |
| X0102 | 30s | M | No | 8 | 32.41 | 34.36 | - | - | 5.49 |
| X0103 | 30s | M | No | 10 | 30.35 | 30.26 | - | - | - |
| X0104 | 40s | F | No | 7 | 21.4 | 21.23 | - | - | - |
| X0107 | 40s | F | No | 9 | 25.17 | 25.01 | - | - | - |
| X0108 | 50s | M | Diabetes mellitus, Hypertension | 6 | 28.57 | 28.89 | - | - | - |
| X0111 | 40s | M | No | 7 | 24.34 | 24.25 | - | - | - |
| X0112 | 30s | M | No | 6 | 31.55 | 32.27 | - | - | - |
| X0113 | 30s | F | No | 7 | 21.8 | 21.67 | - | + | - |
| X0114 | 20s | M | No | 9 | 21.66 | 21.8 | - | - | - |
| X0115 | 30s | M | No | 5 | 26.38 | 26.35 | - | - | - |
| X0116 | 30s | M | No | 9 | 23.53 | 22.89 | - | - | - |
| X0117 | 40s | F | Diabetes mellitus, Hypertension | 6 | 31.61 | 32.47 | - | - | 3.02 |
| X0118 | 40s | M | Hypertension | 6 | 34.09 | 34.9 | - | + | N/A |
| X0119 | 70s | M | No | 18 | 31.15 | 31.3 | - | - | N/A |
| X0120 | 30s | M | No | 6 | 19.69 | 19.58 | - | - | - |
| X0121 | 40s | M | Hypertension | 18 | 21.52 | 21.29 | - | - | - |
| X0122 | 40s | M | Hypertension, | 19 | 34.24 | 34.17 | - | + | 6.71 |
| X0123 | 40s | F | Hypertension, Hypothyroidism | 7 | 23.43 | 23.48 | - | - | - |
| X0124 | 30s | F | No | 11 | 22.69 | 22.36 | - | - | - |
| X0125 | 20s | F | No | 8 | - | - | - | - | - |
| X0129 | 20s | F | No | 6 | - | - | - | - | - |
| X0134 | 30s | M | No | 18 | 27.05 | 26.89 | - | - | - |
| X0135 | 40s | M | No | 1 | 31.99 | 32.54 | - | - | - |
| X0137 | 40s | M | No |  | 25.95 | 25.82 | - | - | - |
| X0139 | 30s | M | No | 31 | - | - | - | - | - |
| X0140 | 30s | F | No | 7 | 24.22 | 24.13 | - | - | - |
| X0141 | 30s | F | No | 7 | 18.71 | 18.68 | - | - | - |
| X0144 | 30s | F | No | 7 | 22.9 | 22.42 | - | - | - |
| X0147 | 40s | F | Hypertension | 8 | 24.36 | 24.33 | - | - | - |
| X0148 | 30s | F | No | 14 | 31.42 | 32.5 | - | - | - |
| X0149 | 50s | F | Hypertension, Hypocholesterolemia | 7 | 35.55 | 34.74 | - | + | 1.9 |
| X0211 | 40s | F | Diabetes mellitus | 10 | 34.23 | 35.89 | N/A | N/A | N/A |
| X0219 | 50s | M | Hypertension | 16 | - | - | N/A | N/A | N/A |
| X0228 | 20s | M | No | 10 | 34 | 33 | N/A | N/A | N/A |

Footnote: NA – Not Available. Co-morbidities documented are listed.

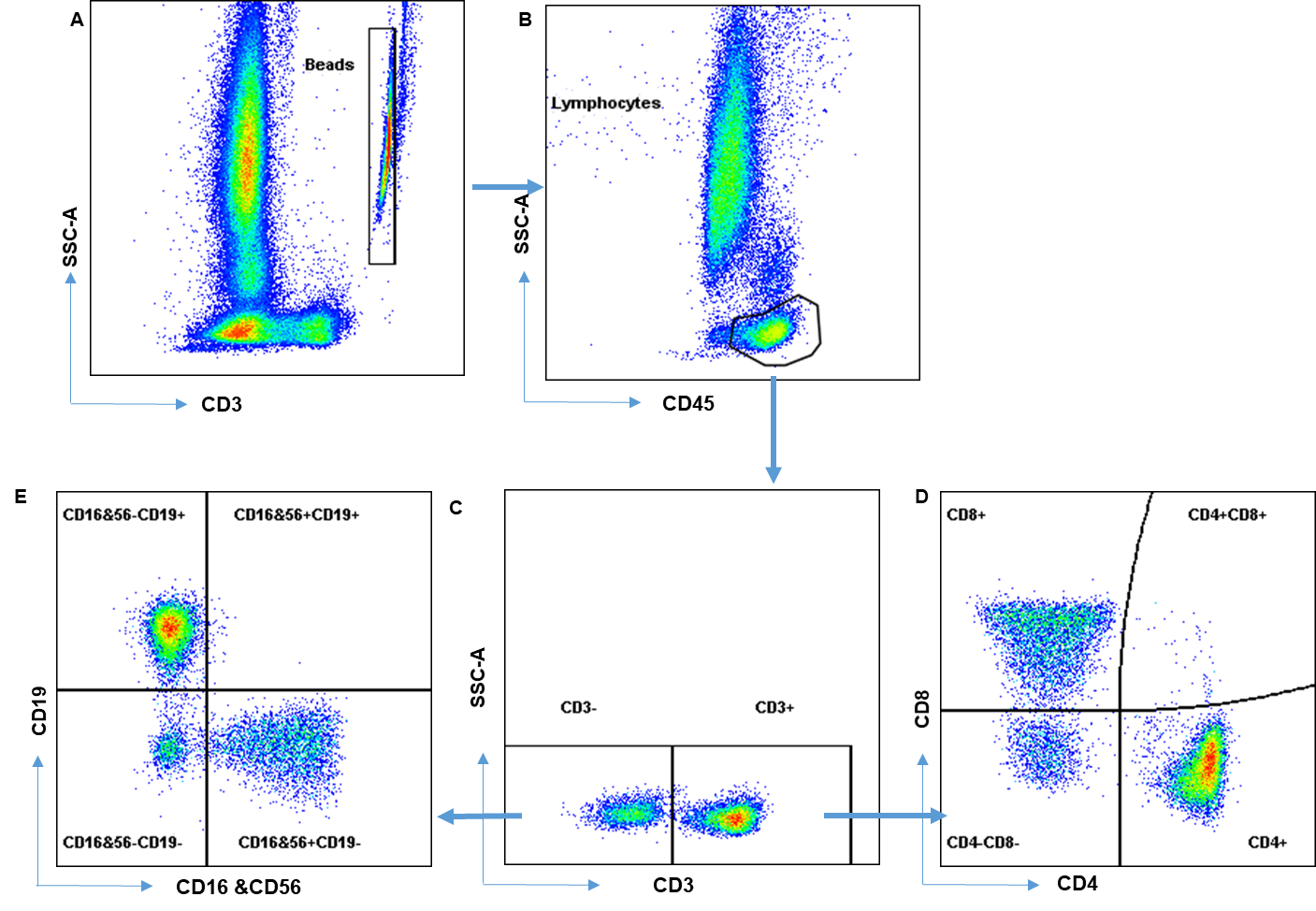

**Supplementary figure S1: Flow cytometric gating strategy for absolute count of T, B and NK cells.** Beads were first identified using CD3 versus SSC-A scatter plot(A). Next, CD45 versus SSC-A scatter plot was used to identify lymphocyte scatter (B). T cells were identified on the basis of expression of CD3 (C) and CD4+ and CD8+ T cells were identified based on CD4 and CD8 expression, respectively (D). B cells and NK cells were identified on the basis of expression of CD19 and CD16 & CD56, respectively.

**(D)**

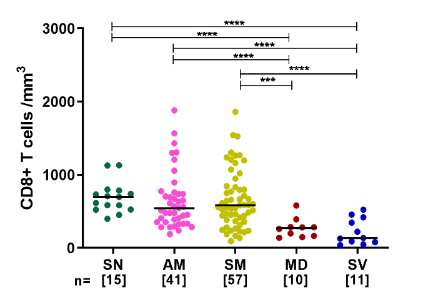

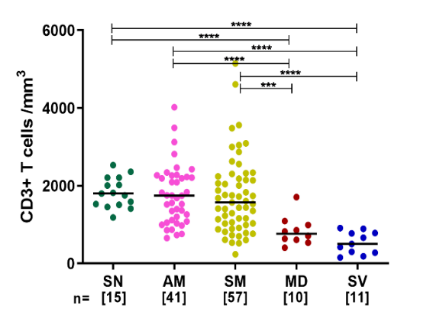

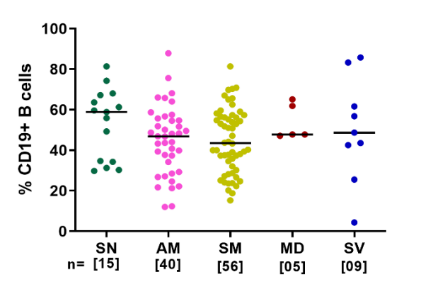

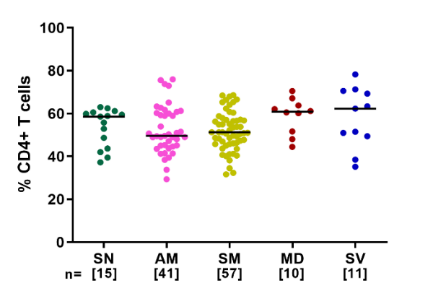

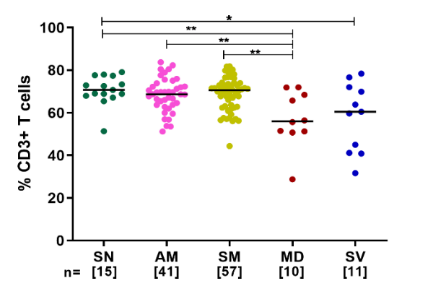

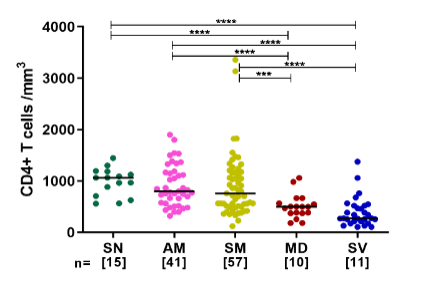

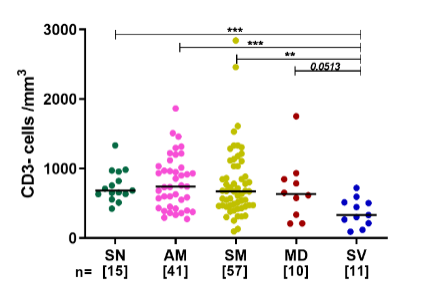

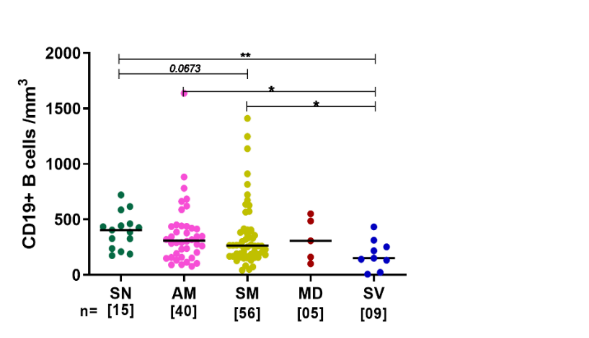

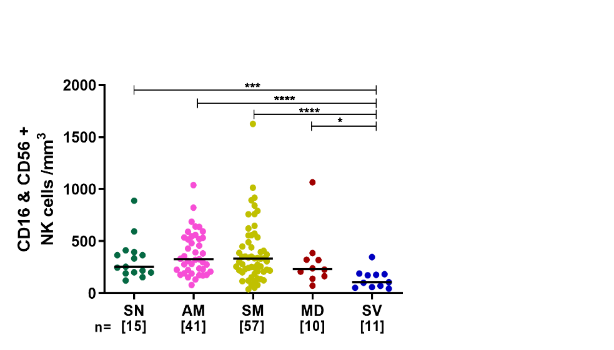

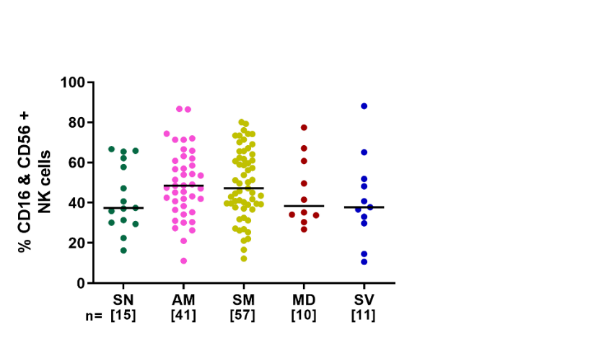

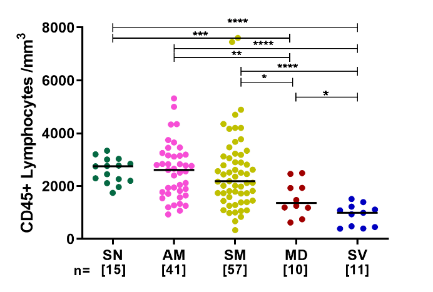

**(B)**

**(C)**

**(A)**

**(F)**

**(G)**

**(E)**

**(I)**

**(H)**

**(M)**

**(L)**

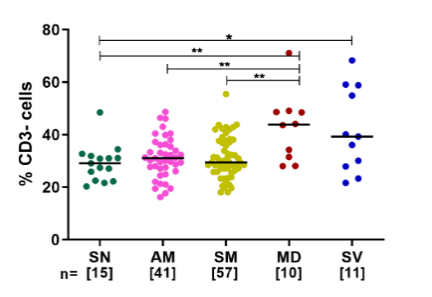

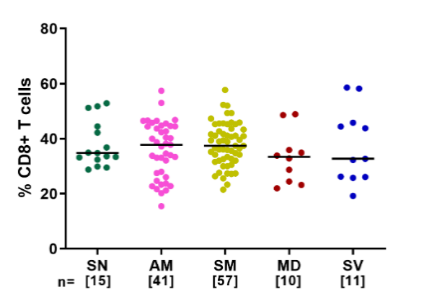

**(J)**

**(K)**

**Supplementary figure S2: Absolute numbers and frequency distribution of T lymphocytes, CD4+ and CD8+ T cells, Non-T lymphocytes, B cells and NK cells among study participants**. **(A-G)** Variation across seronegative (SN), asymptomatic mild (AM), symptomatic mild (SM), moderate (MD) and severe (SV) groups in counts of (A) CD45+ Total Lymphocytes (B) CD3+ T cells, (C) CD4+ T cells, (D) CD8+ T cells, (E) Non-T lymphocytes (CD3- T cells), (F) CD19+ B cells and (G) CD16+& CD56+ NK cells. **(H-M)** Variation across different study groups in frequency of (H) CD3+ T cells (I) CD4+ T cells (J) CD8+ T cells (K) CD3- T cells (L) CD19+ B cells and (M) CD16+& CD56+ NK cells. Statistical significance was calculated by Mann-Whitney U-test; *, p < 0.05; **, p < 0.01, ***, p<0.001 ; and ****, p<0.001.

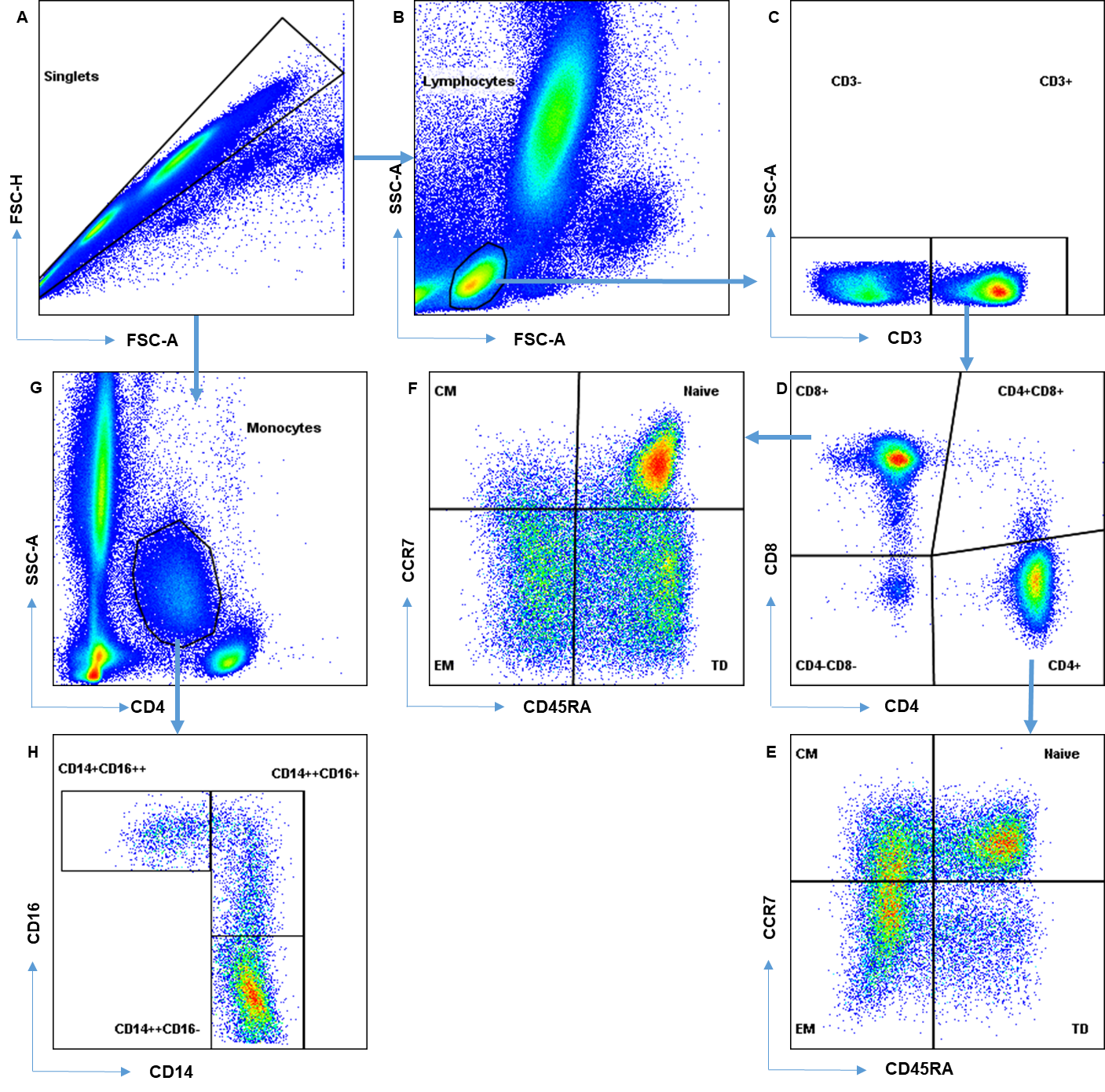

**Supplementary figure S3:** **Flow cytometric gating strategy for distribution of CD4+, CD8+ T cells and monocytes into their functionally distinct subsets.** Singlets were first identified using FSC-A versus FSC-H scatter plot(A) . Next, FSC-A versus SSC-A scatter plot was used to identify lymphocyte scatter (B). T cells were identified on the basis of expression of CD3 (C) and CD4+ and CD8+ T cells were identified based on CD4 and CD8 expression, respectively (D). CD4+ T cell population (E) and CD8+ T cell population (F) was distinguished into naïve (N), central memory (CM), effector memory (EM) and Terminally differentiated (TD) T cell subsets on the basis of surface expression of CD45RA and CCR7. Monocyte scatter was identified using CD4 versus SSC-A (G). Surface expression of CD14 and CD16 on monocytes was used to identify Classical (CD14++CD16-), Intermediate (CD14++CD16+) and Non-classical (CD14+CD16++) monocyte subsets (H).

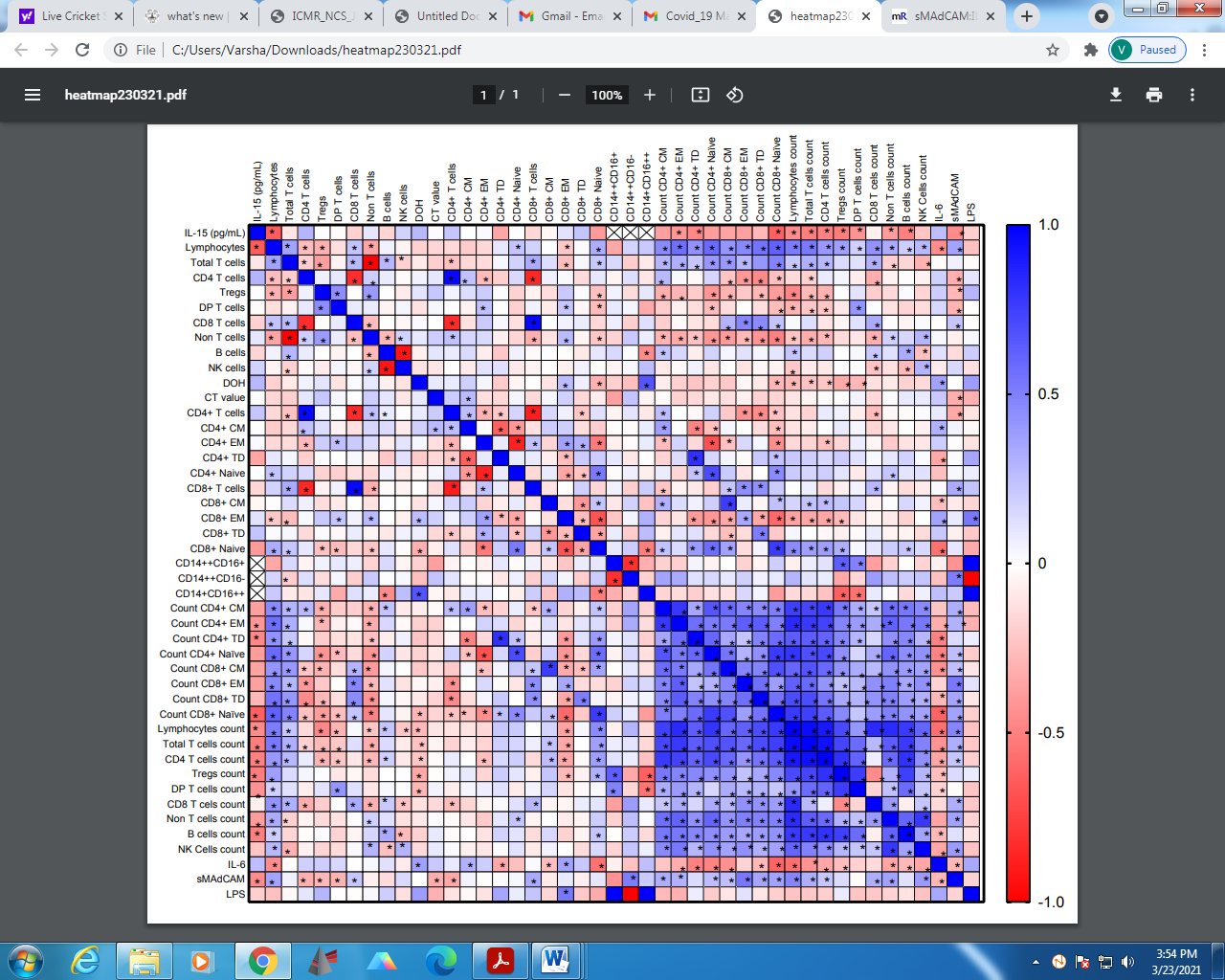

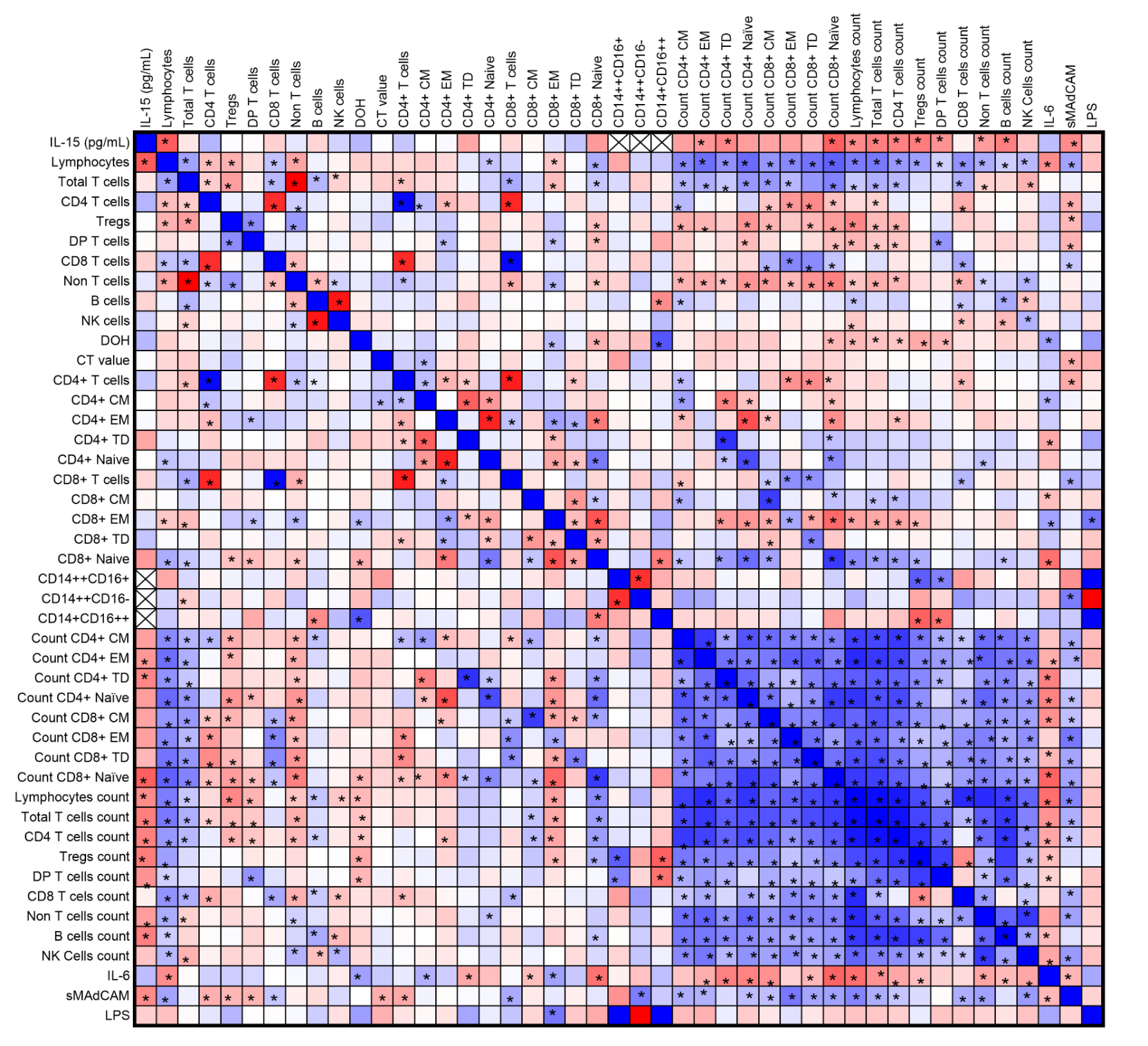

**Supplementary figure S4:** **Correlation matrix of frequency and count of different immune subsets along with IL-6, IL-15 and sMAdCAM in study participants.** The correlation heatmap represents pairwise Spearman correlation matrix of frequencies and absolute count of different immune subset and plasma markers. Red indicates negative correlation and blue indicates positive correlation. Strength of correlation is expressed as a function of colour intensity. Cross indicates absence of enough samples to perform correlation analysis. Correlation analysis was performed using non parametric Spearman Rank Correlation test. *p<0.05 Star indicates statistically significant correlations.

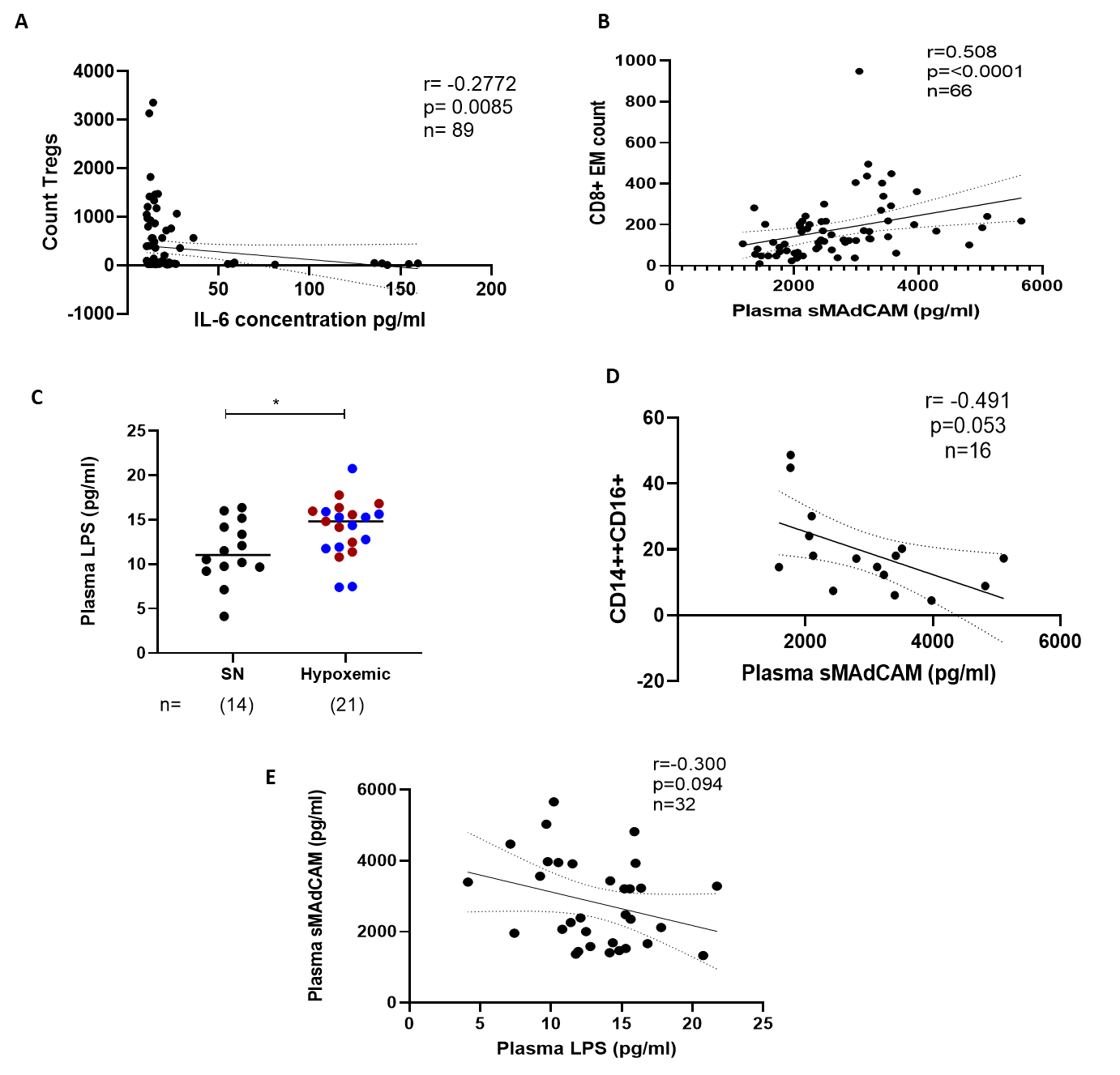

**Supplementary figure S5**: Association of (A) IL-6 with Treg count and (B) sMAdCAM with Intermediate monocyte frequency. (C) Plasma LPS levels among study participants. (D) Association of sMAdCAM with intermediate monocytes. (E) Association of Plasma LPS with sMAdCAM. Statistical significance was calculated by Mann-Whitney U-test, * p < 0.05. Correlation analysis was performed using non parametric Spearman Rank Correlation test.
